# AB_x_Sure for differentiating bacterial from viral Infection

**DOI:** 10.64898/2026.03.09.26347921

**Authors:** Neha Khaware, Rishikant Thakur, Kartik Kachhawa, Prabhu Balasubramanian, Vivekanandan Perumal, Ravikrishnan Elangovan

## Abstract

Bacterial and Viral infections have identical symptoms, making it difficult to diagnose based on visible symptoms. This results in the overprescription of antibiotics, even in cases of viral infections, to avoid missing bacterial infections that could progress into a more severe condition and/or sepsis. This diagnostic gap contributes to the overuse of antimicrobials. To address the issue, a deployable diagnostic test has been developed and evaluated that can help differentiate between bacterial and viral infections. The approach is centred on three crucial biological markers: CD64 expression on leukocytes to detect bacterial infections, CD169 expression on leukocytes to detect viral infections and total leukocyte count as a general health indicator of the patient. AB_x_Sure comprises a cartridge for sample processing, a device with in-built pneumatics, and an optical reader for quantifying biomarkers from blood. The AB_x_Sure performance was tested and compared with a commercial plate reader and flow cytometer, yielding a promising correlation of 0.89. This comprehensive test will potentially provide clinicians with valuable information for effective and timely treatment.

## 1. Introduction

Identifying infections often involves examining blood components and hematological indicators such as neutrophil, WBC, RBC, monocyte, lymphocyte, and platelet counts, as well as erythrocyte sedimentation rate. However, these markers lack specificity and cannot differentiate between various infection types, thus limiting their diagnostic usefulness. While blood culture tests are considered the best method for confirming bacterial infections, they have drawbacks, including reduced sensitivity due to previous antibiotic use, sampling errors, and delayed results. The 24 to 48-hour time required for blood cultures makes them impractical in situations demanding quick results, often leading to prescribing antimicrobials based solely on symptoms (1,2).

Common symptoms such as fever, fatigue, and coughing often overlap between bacterial and viral infections, complicating the diagnostic process (3). This leads to the overprescription of antibiotics even in cases of a viral infection (Figure 1). Misuse and overuse of antimicrobials are the significant drivers behind AMR development. Bacterial infections, particularly antibiotic-resistant bacteria, were responsible for about 4.5 million deaths in 2019 and are projected to cause approximately 10 million deaths by 2050 (4). Therefore, a diagnostic test that can efficiently detect the pathogen or a specific immune response is the need of the hour.

**Figure 1:**
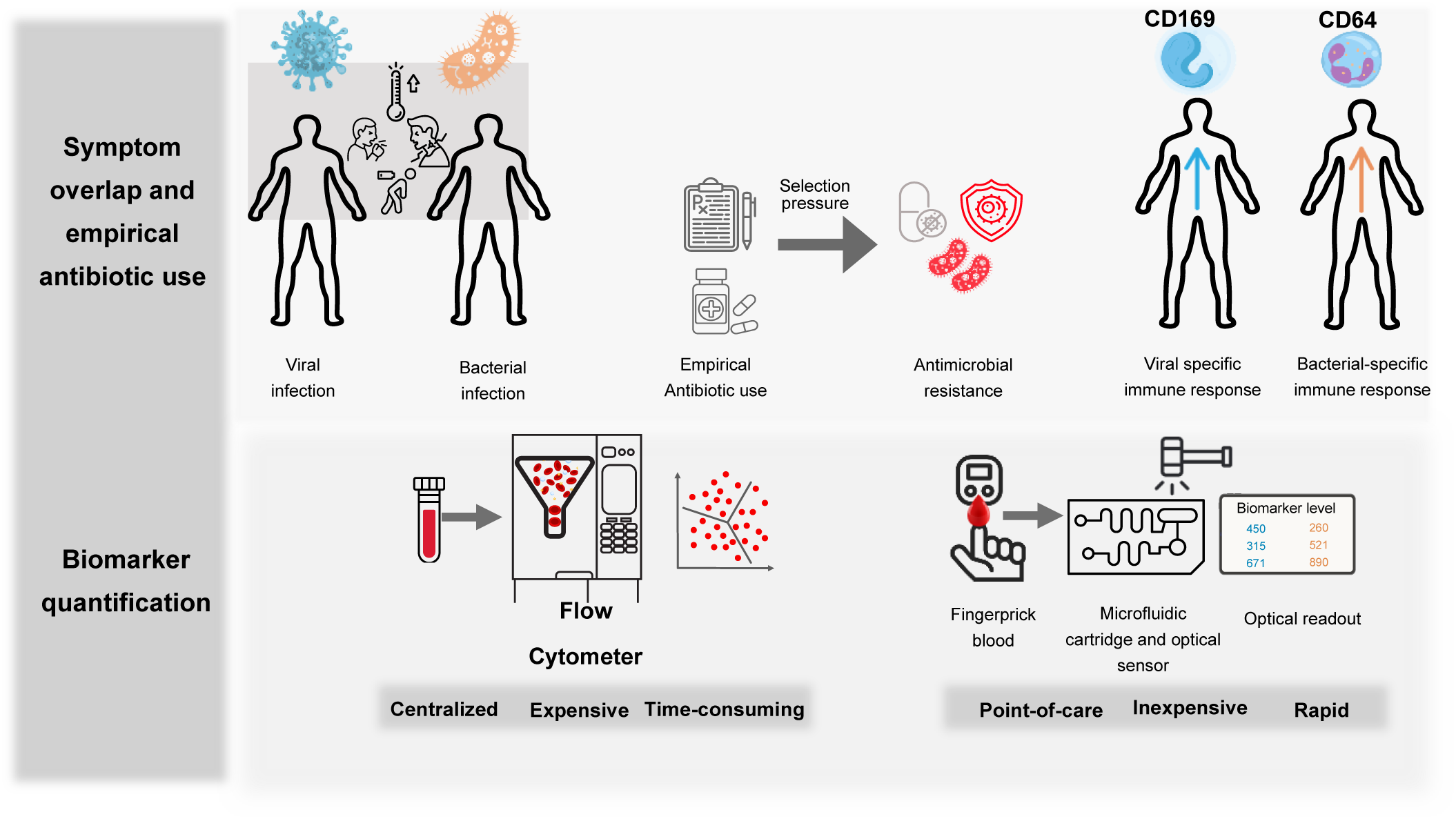
Workflow illustrating symptom overlap, antibiotic overuse, and immune biomarker screening. Overlapping symptoms between viral and bacterial infections drive empirical antibiotic prescribing and contribute to antimicrobial resistance. Although host immune biomarkers can distinguish infection etiology, their measurement typically requires centralized flow cytometry. The proposed microfluidic–optical screening device enables rapid, quantitative immune biomarker assessment from a finger-prick blood sample, supporting point-of-care differentiation of viral and bacterial infections.

The current diagnosis relies on the use of microbiological and nucleic acid-based tests (RT-PCR) to detect the pathogen but has its challenges, such as requiring a longer time to produce results and the presence of the pathogen at an inaccessible location (pneumonia). Therefore, a different diagnostic approach is required to overcome these challenges. Monitoring the patient’s immune response rather than directly detecting the pathogen has the required potential. Circulating host proteins can be rapidly measured and is a promising source of biomarkers for bacterial and viral infections. Currently, various proteins are evaluated in routine clinical practice, including C-reactive protein, Procalcitonin, and Interleukin-6, to determine the infection type. However, their utility in differentiating bacterial from viral infection and inflammation is limited (5).

CD64 (FcγRI), an Fc receptor for IgG, is naturally expressed on macrophages, monocytes, and eosinophils with minimal presence on resting neutrophils. However, the expression of neutrophil CD64 (nCD64) significantly increases when neutrophils are activated in response to microbial wall components, complement split products, and certain pro-inflammatory cytokines such as IFN-γ and granulocyte colony-stimulating factor (G-CSF) (6–9). Neutrophil CD64 expression on resting neutrophils is relatively low, at approximately 1000 molecules per cell. However, in severe infections, its expression can increase 8- to 10-fold. This upregulation occurs within 4–6 hours of infection onset, making it an early-phase biomarker. This rapid response highlights its critical role as a marker for bacterial infection, inflammation, and sepsis (10,11). CD169 is a member of the sialic acid binding immunoglobulin-like lectins (SIGLEC) family. It is an endocytic receptor that binds to sialylated bacteria and viruses. In response to interferon α (type 1 interferon) in viral infection, the expression of CD169 increases exponentially on the surface of monocytes (12,13). The role of CD169+ monocytes has been well studied in viral infections, especially human immunodeficiency virus (HIV), respiratory syncytial virus (RSV), and, more recently, SARS-CoV-2 (14). The combination of CD64 and CD169 biomarkers can help differentiate bacterial from viral infections, thereby aiding better diagnosis and treatment. CD64 and CD169 have been evaluated for their ability to differentiate between bacterial and viral infections, with ROC-AUC of 0.846 and 0.841, respectively (13). These biomarkers are currently assessed using flow cytometry, a time-consuming and costly method. Flow cytometry tests can cost up to $30 per test and analyze only 10-15 samples per day, which makes it unsuitable for widespread use in clinical settings (15). Therefore, there is a growing need for a point-of-care (PoC) device that can provide rapid, cost-effective, and easily accessible infection diagnosis. The development of such a device would address current diagnostic challenges and improve patient care (16).

In this work, CD64 and CD169 biomarkers expressed on leukocytes have been explored to develop an assay for distinguishing bacterial from viral infections. CD64 is used in combination with CD169. An assay for rapid and accurate quantification of these markers is developed and integrated into a microfluidic device called AB_x_Sure. Along with these biomarkers, total white blood cells are also quantified in AB_x_Sure to provide an all-in-one infection diagnosis system. AB_x_Sure consists of a microfluidic cartridge for sample processing and a device with an in-built optical reader for liquid handling. The blood cells are lysed and labeled with fluorophore-conjugated antibodies to quantify CD64 and CD169 and a nucleic acid-binding fluorescence dye for total leukocyte count.

## 2. Methods and Materials-

The system integrates a microfluidic cartridge for liquid handling and a device for controlling pneumatics, automation, and fluorescence detection, as illustrated in Figure 2. The cartridge consists of a sample chamber, a wash chamber, three sample wells, a mixing loop, three membranes (for cell capture), and a dump chamber. Six inlets/suction points were incorporated to control the liquid flow, and the inner channel dimensions are 0.8mm. The wash and sample chamber, with a volume capacity of 720 μL, ends in a common channel to allow streamlined fluid flow into the sample wells. The sample was introduced through a syringe into the sample chamber, from which it moved to the sample wells, one after the other, filling each well with 100 μL of sample. Each sample well is connected to a mixing loop (252mm long) to ensure an even mixing of the sample with the dye. The mixing was enabled by back-and-forth motion facilitated by the peristaltic pump. After mixing, the sample moves to the membrane for the cell enrichment, and the remaining liquid moves to the dump ahead of the membrane, which can hold up to 1.35mL liquid. The cartridge is inserted into the device casing using a slot-based alignment mechanism. The optical reader then reads the membranes. The device consists of a touchscreen pad that allows users to control the system with the help of a Graphical User Interface (GUI).

**Figure 2:**
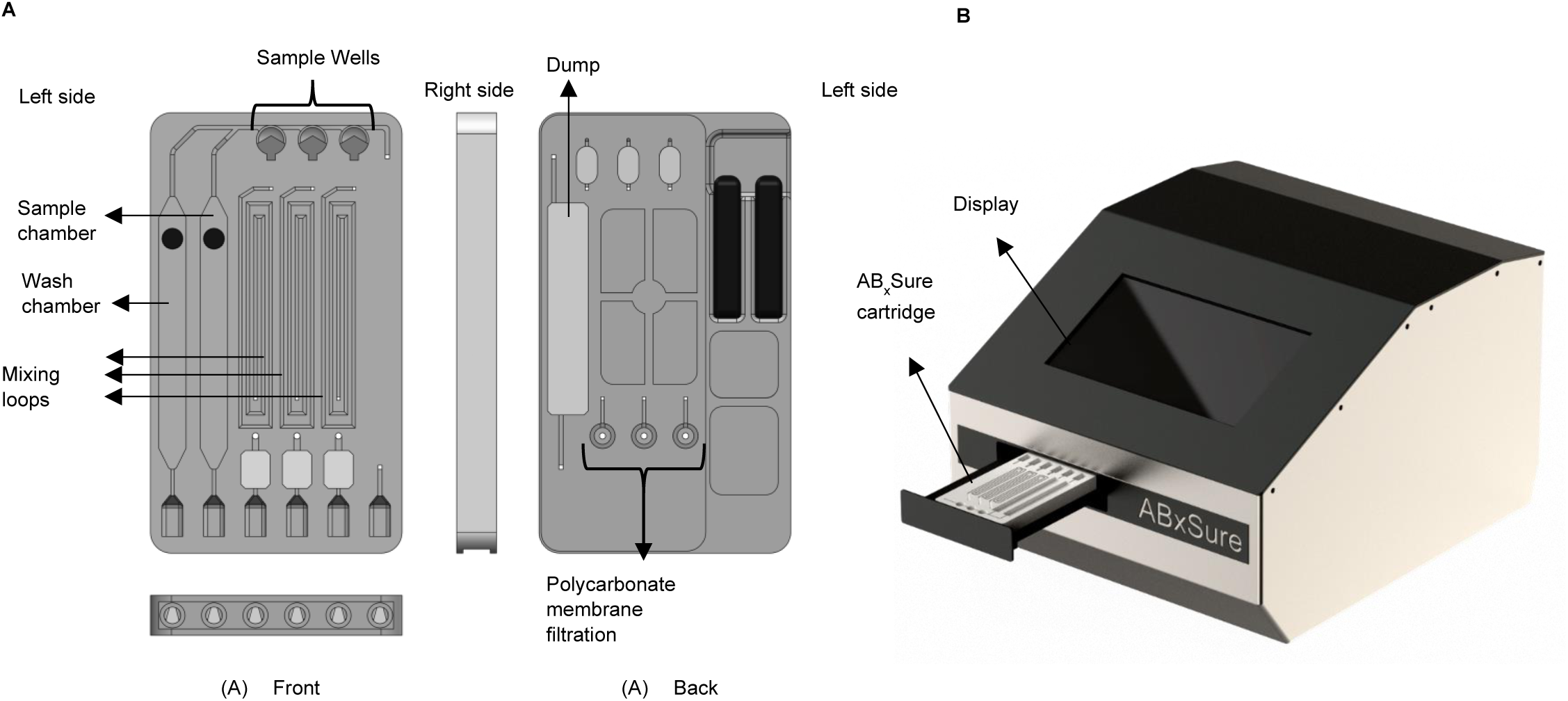
Microfluidic cartridge and integrated optical screening device for immune biomarker quantification. A. Labeled front and back views of the microfluidic cartridge. B. Integrated screening device illustrating cartridge insertion, pneumatic module and motorized actuation, optical detection module, and display for quantitative biomarker readout.

The cartridge was fabricated using stereolithography (SLA)-based 3-D printing with a ProJet MJP 2500 Plus, using VisiJet M2R-CL and M2 SUP (CL=clear and SUP=support) materials, designed in SolidWorks software. Figure 2 shows the microfluidic cartridge with dimensions of 90 × 56 × 9mm (L × B × H). Polypropylene (PP) was used as the cartridge assembly material, providing mechanical stability and compatibility with biological samples. The membrane was laser cut into a circular shape with a diameter of 5.5mm. The membranes were placed in a designated hollow cavity in the cartridge and secured with adhesive to ensure leak-free operation. For sterilization and cleaning, distilled water was used for boiling procedures, while a 70–90% ethanol spray was applied for surface disinfection. Drying of components was facilitated by a compressed air gun. A sponge material was included to serve as a dump zone for absorbing excess fluid within the cartridge. To ensure robust sealing and adhesion, a double-sided industrial-grade tape was used. Finally, pre-cut polycarbonate (PC) sheets, fabricated via laser cutting, were used as transparent covers to seal the membrane trench or optical window.

### 2.1 AB_x_Sure design and operational protocol

The system can quantify three biological markers for diagnosing infection, effectively distinguishing between bacterial and viral infections. The device requires no manual steps, making it accessible to users with little or no expertise. The cartridge is preloaded with reagents for efficient blood sample processing. The sample chamber contains an RBC lysis buffer that lyses whole blood, eliminating background interference and reducing viscosity, enabling smooth sample flow. The sample is then directed to the sample well, where it is evenly split into three wells for simultaneous quantification of two surface antigens and total cell count. This unique capability allows users to analyze multiple biomarkers in a single run. In the labeling and mixing loop, the blood cells are precisely labeled with immobilized antibodies or dye. This minimizes manual intervention and extends reagent shelf life. Pneumatic pumps facilitate continuous back-and-forth mixing, ensuring thorough sample incubation and optimal labelling (Supplementary Figure S3). The processed sample is subsequently captured on a polycarbonate membrane (pore size of 3μm), designed to retain all blood cells. After capture, the membranes are washed with the preloaded wash buffer in the wash buffer chamber. Finally, the membrane is analyzed using an optical detector that delivers results in relative fluorescence units.

#### 2.1.1 AB_x_Sure Working

The sample chamber pre-loaded with 450μL of RBC lysis buffer into which 50μL of raw sample is introduced and inserted into the device. After a 5-minute incubation to ensure complete lysis, the lysate is automatically directed into the sample reaction wells. The fluidic design ensures precise distribution of 100 μL into each well, while excess volume is diverted to a dedicated dump reservoir. This reservoir incorporates an absorbent pad to provide a capillary draw and effectively prevent backflow into the active channels. Within each sample well, the sample enters a specialized mixing loop containing the immobilized antibodies or the dye. The samples move into the mixing loop in a back-and-forth motion for 15 minutes to facilitate the mixing and incubation of the cells with the antibody and the dye. After mixing, the sample is transitioned to the membrane where the cells are captured and the residual liquid is collected in the chamber ahead of the membrane. The membrane is a polycarbonate track-etched with a pore size of 3μm, carefully laser cut into a 5.5mm diameter. Similarly, the wash buffer moves from the wash chamber to the sample well, to the mixing loop, and to the membrane, removing the unbound dye or antibody. This reduces the background fluorescence. The optical reader now reads the cartridge, which reads all three membranes one by one and displays the readings in the form of relative fluorescence units.

#### 2.1.2 AB_x_Sure Experimental protocol

The sample was prepared by mixing 100μL of whole blood with 900μL of RBC lysis buffer (1:10). The sample was divided into three parts (250μL each) and labelling antibody or dye was added in appropriate concentrations. 100μL of prepared samples were added to all three collection wells directly. A suction pressure was applied from the bottom of the cartridge (each well has a separate mixing loop, membrane and suction points). The cells were captured on the membrane. The wash buffer (1XPBS) was added to the collection well, and again, suction pressure was applied. This was done to remove the unbound antibody/ dye. The membranes were then read in the AB_x_Sure optical reader and readings were recorded.

#### 2.1.3 AB_x_Sure Electronic assembly

The electronics of AB_x_Sure ensures a fully automatic liquid control, data acquisition and display. An external 26V power supply powered the device via an adapter, protected by an 8A fuse for overcurrent protection. Multiple DC–DC converters were used to generate regulated voltages for individual subsystems, ensuring stable operation and electrical isolation. The hardware has been designed modularly where the microcontroller, an integrated circuit interacts with the GUI via touchscreen pad and feedback pressure sensor (helps adjust the flow rate and pressure withing the channels) to initiate the program. It controls the LED, which excites the sample, and the photodetector, which collects the fluorescence signals, and the analog digital converter (ADC) converts these signals into relative fluorescence units and is displayed on the screen. It also controls the liquid flow rate using the peristaltic pump. The wash and sample chamber require positive pressure to move the liquid further to the sample collection well and finally to the dump, which requires negative pressure (suction). The mixing loops require both positive pressure and suction for the back-and-forth motion. The feedback pressure sensor controls all the liquid flow and communicates with the microcontroller.

### 2.2 Methodology

#### 2.2.1 In Vitro Validation of Biomarker Expression

To establish the biological responsiveness of the selected surface antigens, Human Leukemia 60 (HL60) cells were utilized as models for neutrophils and monocytes. For neutrophils, the HL60 cells were incubated with 1.3% DMSO for 8 days and for monocytes, the HL60 cells were incubated with 2ng/mL of phorbol myristate acetate (PMA) (Supplementary Figure S1). Cells were stimulated with 1600 IU/mL Interferon-gamma (IFN-γ) and 50ng/mL interferon alpha (IFN-α) to simulate bacterial (CD64) and viral (CD169) signalling environments, respectively. CD64 and CD169 expression levels in uninduced vs. induced populations were quantified using a commercial microplate reader to define the baseline expression range for each biomarker (Supplementary Figure S2).

#### 2.2.2 WBC calibration in AB_x_Sure

The clinical samples (leftover whole blood) were acquired from the IITD hospital pathology laboratory, Indian Institute of Technology, Delhi, New Delhi, India. The required ethical clearance for performing the study was obtained and all the procedures were followed as per the guidelines of the Institute Ethics Committee, IIT Delhi (Ethical clearance reference number-Ethics application 2024/P043-21-03-2024). About 100 μL of whole blood was lysed with 1X lysis buffer [349202, BD Pharmingen, USA (www.bdbiosciences.com)] (900 μL) for 5 minutes, and then centrifuged at 300 × g for 8 minutes. The WBC pellet was resuspended in 500μL of 1X PBS [P4417-100TAB, Sigma] buffer. 20μL of cells were stained with methylene blue and counted in a hemocytometer. After the cell count, the dilutions ranged from 10^3^ cells per mL to 10^6^ cells per mL. Each dilution was labeled with 5μM Syto85 dye [S11366, ThermoFisher Scientific USA]. The sample was incubated for 10 minutes in the dark. 500μL of the sample was loaded into the sample chamber in the cartridge in triplicates.

#### 2.2.3 CD64-CD169 quantification

CD64 and CD169 markers were quantified in the multimode plate reader, AB_x_Sure, and flow cytometer. This was done to ensure that AB_x_Sure results are comparable with those of the commercial system and the gold standard.

For the proof of concept, the Human Leukemia 60 (HL60) cell line was used as the source of in-vitro neutrophils and monocytes. For neutrophils, the HL60 cells were incubated with 1.3% DMSO for 8 days; the media was changed every three days (Figure 4.1). The differentiated HL60 cells were observed under the microscope to confirm. For monocytes, the HL60 cells were incubated with 2ng/mL of phorbol myristate acetate (PMA) (different concentrations of PMA were evaluated for the differentiation-1ng/mL to 10 ng/mL) for 2 days (Figure 4.2). All the incubation was done in the CO2 incubator at 37oC and 5% CO2.

**Figure 3:**
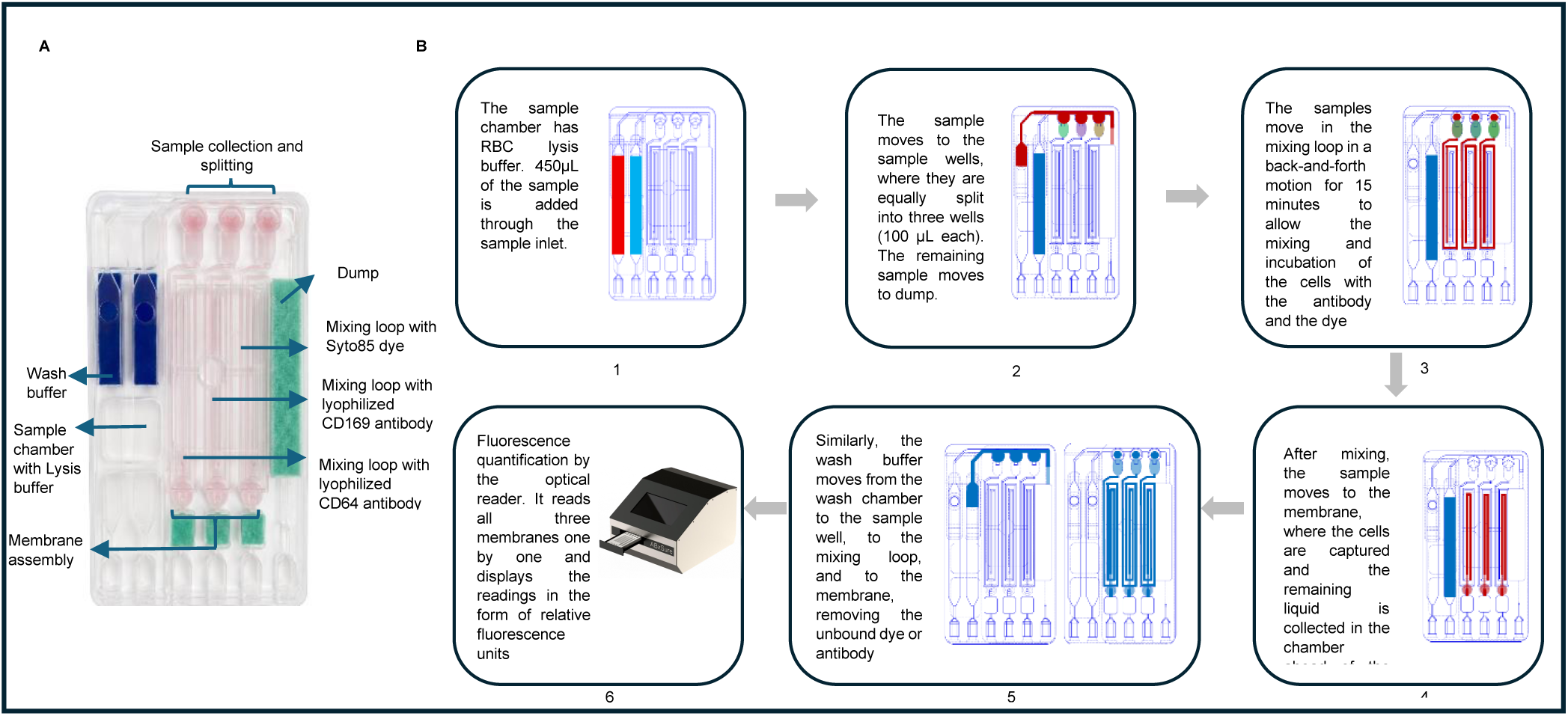
A. Front side of the assembled cartridge, B. Schematic representation of the ABxSure workflow. (1) Sample addition and RBC lysis; (2) Equal distribution into the sample wells; (3) On-chip labeling via serpentine mixing; (4) Size-based cell separation on the membranes; (5) Washing of the residual/ unbound dye or antibody; (6) In-situ quantification of the captured cellular fraction.

**Figure 4:**
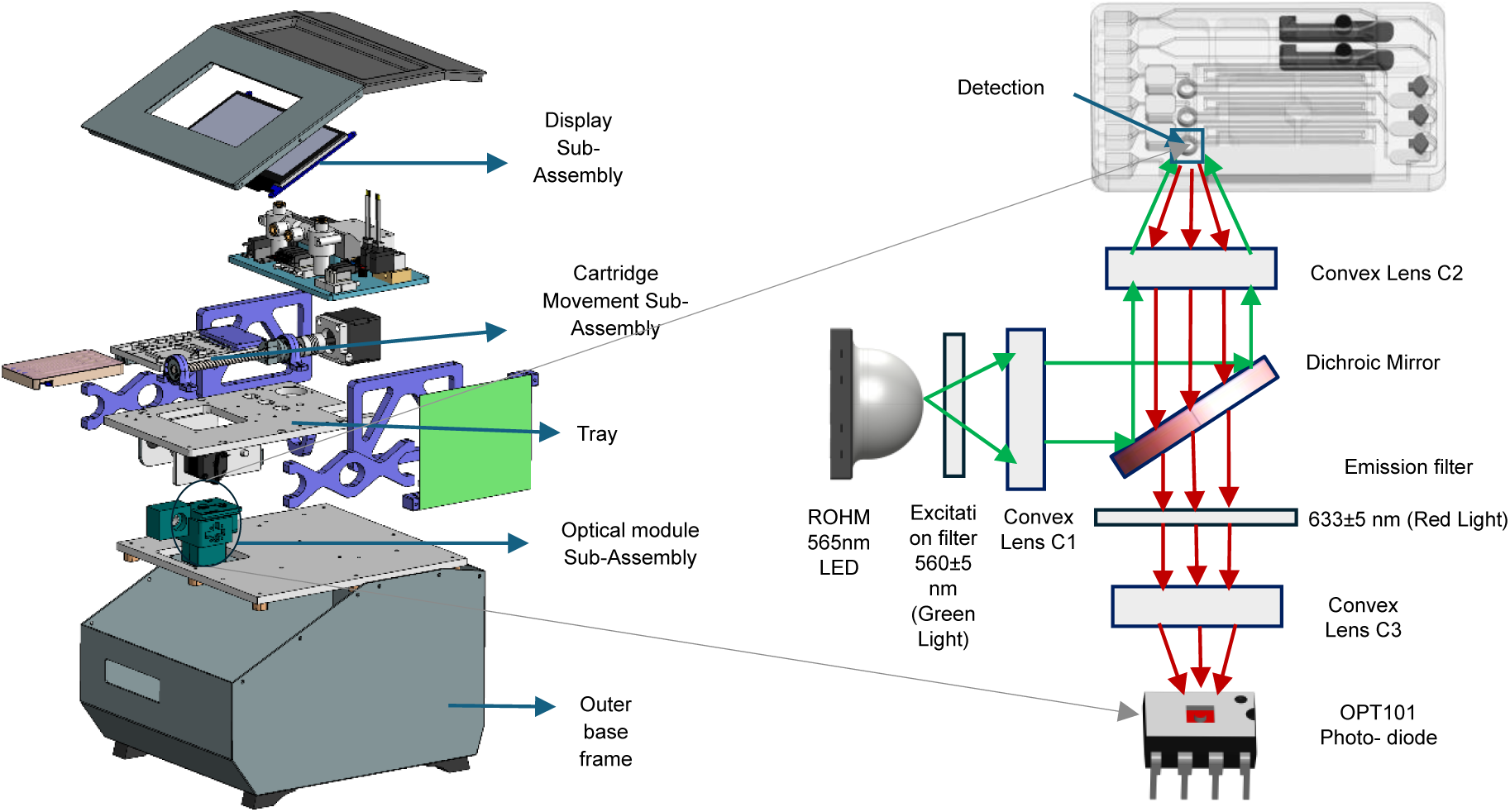
Schematic representation of the integrated microfluidic detection system. The device houses the optical module comprising a high-power LED, collimating optics and a photodiode aligned with the cartridge interface

#### 2.2.4 CD64-CD169 quantification in plate reader

In 50μL of whole blood, 450μL of 1X lysis buffer [349202, BD Pharmingen, USA (www.bdbiosciences.com)]. was added and incubated for 5 minutes. The sample was then centrifuged at 300 x g for 8 minutes to pellet down the WBCs. The WBCs were counted in a hemocytometer [Rohem, India], and dilutions were prepared in 1XPBS ranging from 10^3^ to 10^6^ cells/mL.

Thereafter, 3μL of CD64-PE [558592, BD Pharmingen, USA (www.bdbiosciences.com)] or 1μL of CD169-PE [565248, BD Pharmingen, USA (www.bdbiosciences.com)] antibody was added to all the dilutions and incubated for 25 minutes in the dark. After incubation, the cells were again centrifuged at 300 x g for 8 minutes to remove the unbound dye. The samples were then added to the 96-well plate, and readings were recorded in the multimode plate reader at ex/em of 530/625nm.

#### 2.2.5 CD64-CD169 quantification in AB_x_Sure

In 50μL of whole blood, 3μL of CD64-PE or 1μL of CD169-PE antibody was added and incubated for 25 minutes in the dark. After incubation, 450μL of 1X lysis buffer was added and incubated for 5 minutes. The samples were loaded in the AB_x_Sure cartridge in duplicates, as explained earlier in AB_x_Sure experimental protocol (Section 2.1.2). After cell capture, the cartridges were read by the AB_x_Sure optical reader. Then, the membranes were removed from the cartridge and put in the 96-well plate. The plate was also read, and readings were recorded in the multimode plate reader at excitation and emission of 530/625nm.

#### 2.2.6 Flow cytometry protocol

The CD64 and CD169 biomarkers were quantified in the blood samples in three systems-AB_x_Sure, plate reader and flow cytometer. The blood samples after labeling and RBC lysis were examined in the flow cytometer. The quantification of total CD64 and total CD169 expression was done using phycoerythrin (PE) labeled antibodies. A commercial PE quantification kit called the BD Quantibrite Phycoerythrin fluorescence quantification kit [340495, BD Pharmingen, USA (www.bdbiosciences.com)] was used to quantify PE molecules per cell. Each BD Quantibrite PE tube contains a lyophilized pellet of bead conjugated with four distinct levels of phycoerythrin (PE). The number of PE molecules per bead is provided with each kit. These beads are intended for use with PE-labeled monoclonal antibodies to estimate the antibody bound to each cell via flow cytometry. When the beads are analyzed under the same instrument settings as the assay, the FL2 axis can be translated into the number of PE molecules bound to each cell. The PE molecules per cell can be converted into antibodies bound per cell by applying known PE-to-antibody ratios (Supplementary Figure S4).

The blood samples were run after the PE calibration beads in BD Fortessa X20 flow cytometer, and the total CD64 and CD169 antibodies bound per cell were calculated [Geometric mean fluorescence * Events= total CD64/CD169-PE molecules expressed in the sample]. The results were compared among all three systems and correlation was calculated using OriginPro.

## 3. Results

### 3.1 Characterization of Biomarker Responsiveness in Cell-Line Models

Prior to device validation with clinical samples, the differential expression of the targeted biomarkers was confirmed in induced cell-line models. Stimulation with IFN-γ and IFN-α resulted in significant upregulation of the CD64 (bacterial-associated) and CD169 (viral-associated) markers, as confirmed by plate reader analysis (Supplementary Figure S2). This controlled in vitro validation established the sensitivity of our selected biomarkers to cytokine signalling, providing a biological baseline for the subsequent quantification in whole blood samples using the ABxSure.

### 3.2 WBC quantification in AB_x_Sure

The whole blood was lysed and labeled with 5μM Syto85 dye for 10 minutes. The sample was then loaded in the sample chamber and flow was controlled manually using syringes. The sample moved to the sample well, splitting 100μL into each sample well, and the remaining sample went to the dump. In a similar manner, the wash buffer was loaded in the cartridge and membranes were washed to remove the unbound dye. The cartridge was then read in the AB_x_Sure optical reader. For comparison, the membranes were taken out of the cartridge and read in the multimode plate reader. The readings were recorded, and graphs were plotted in Origin software (Figure 5A).

**Figure 5:**
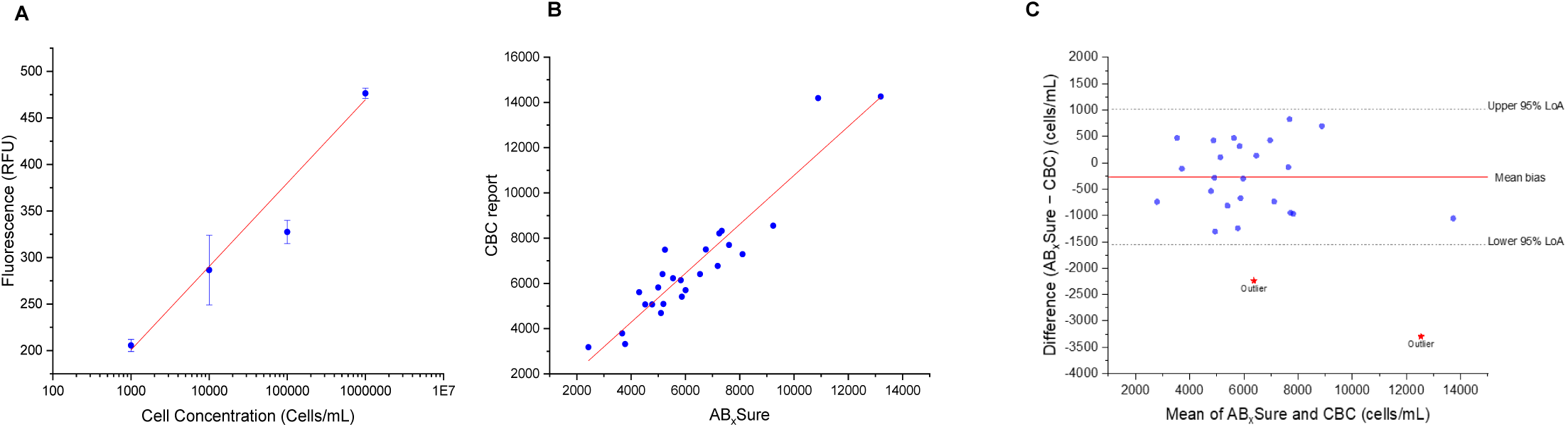
A. WBC calibration plot in ABxSure. The WBCs were labelled with Syto 85 dye. A plot of the fluorescence intensity (y-axis) as a function of cell concentration (x-axis), ranging from [103] to [106] cells/mL. Each data point represents the mean of three independent replicates, with error bars indicating the standard deviation (SD). The solid line indicates the linear-square fit (y = 89.66x – 68.31). **B. WBC count in ABxSure vs CBC report (n=25)**. The solid line indicates the linear-square fit (y = 1081x – 46.03) R2= 0.88, r= 0.93. **C. Bland–Altman plot comparing WBC measurements obtained from the device and CBC analyzer.** The red dot represents outliers.

### 3.2 WBC quantification comparison between AB_x_Sure and cell counter

The calibration curve, established with a known number of white blood cells (WBCs), was employed to quantify the total WBCs in the blood samples. The R^2^ value for the linear regression fit was 0.98. The total WBC count for each blood sample was obtained from the hospital records. A plot of fluorescence intensity versus cell concentration (cells per μL) was generated, and linear regression analysis was performed using OriginPro software. The fluorescence data from the unknown samples were then used to calculate their WBC count (cells/μL). The WBC counts from the hospital and AB_x_Sure were compared for 25 samples, yielding a correlation coefficient of 0.93. This high correlation demonstrates that AB_x_Sure is highly effective in accurately quantifying total WBCs (Figure 5B). A Bland–Altman analysis was used to assess agreement between the device and the CBC analyzer for WBC quantification after exclusion of two outliers. The mean bias was −266 cells/mL, indicating a slight underestimation by the device. The 95% limits of agreement (LoA) ranged from −1551 to +1019 cells/mL, with most measurements falling within these limits. The 95% confidence interval of the mean bias included zero, suggesting no statistically significant systematic difference. No clear proportional bias was observed, although increased variability was noted at lower WBC values.

### 3.3 CD64-CD169 quantification in plate reader vs AB_x_Sure

About 20 whole blood samples were collected from the IITD hospital and were processed as mentioned in the methods section. The readings were recorded in the AB_x_Sure, multimode plate reader [VictorNivo by Revvity] and the flow cytometer. The results from both systems (plate reader and AB_x_Sure) were recorded and plotted in the *OriginPro* software. The x-axis represents AB_x_Sure, and the Y-axis represents the plate reader. The blank fluorescence was subtracted from the readings obtained from both the system and then plotted. The correlation between the two systems was calculated by the software and was found to be 0.899 for measuring CD64 expression and 0.89 for measuring CD169 expression in the blood samples.

Agreement between the AB_x_Sure and the multimode plate reader was evaluated for both CD64 and CD169 biomarkers using log–log calibration followed by log-ratio Bland–Altman analysis. Prior to calibration, systematic differences were observed due to differences in the measurement scale. After calibration, the mean log₁₀ bias was effectively eliminated for both biomarkers, indicating the absence of systematic error. For CD64, the 95% limits of agreement ranged from −0.265 to +0.265 log₁₀ units, corresponding to 0.54- to 1.84-fold relative to the plate reader (Figure 6 B). In contrast, CD169 demonstrated substantially tighter agreement, with 95% limits of agreement from −0.064 to +0.064 log₁₀ units, equivalent to 0.86- to 1.16-fold (Figure 6 F).

**Figure 6:**
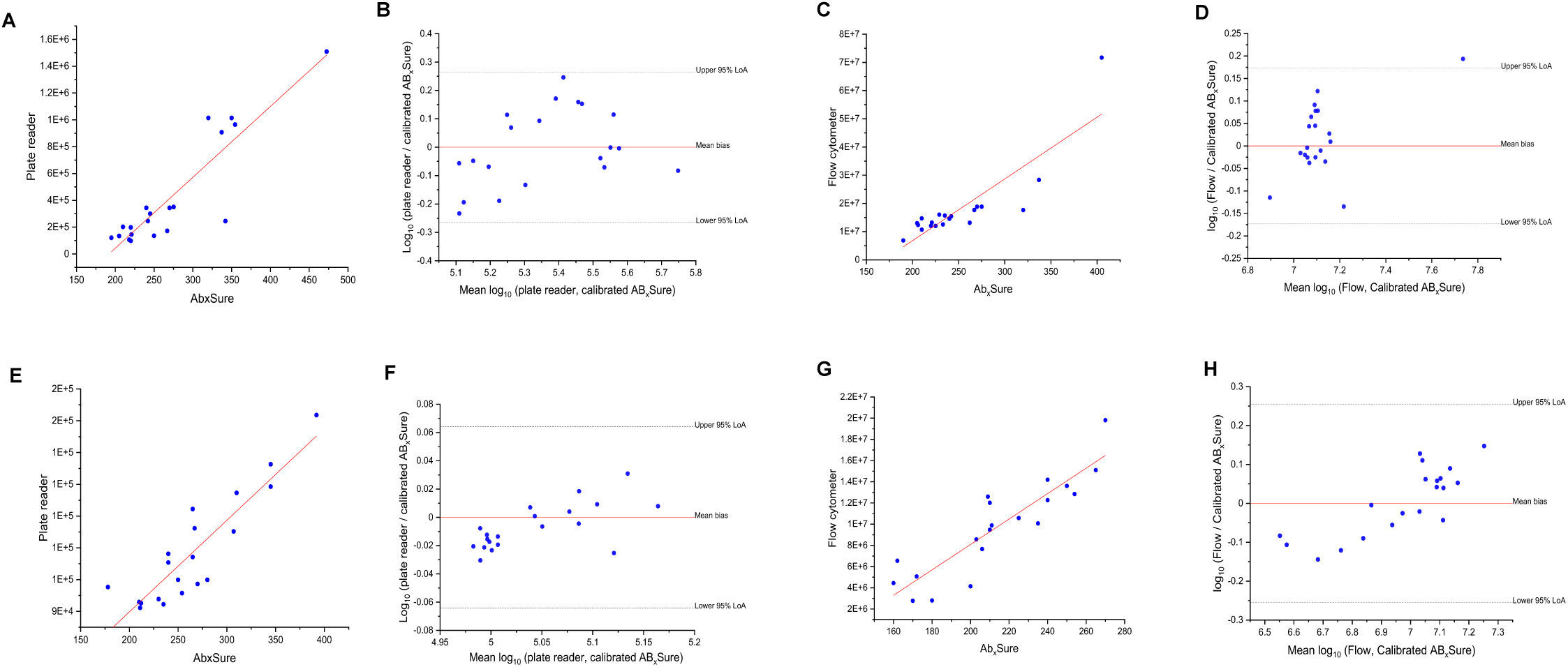
A. CD64 expression quantification in plate reader and ABxSure (n=20). The solid line indicates the linear-square fit (y =5286x – 1014094.9), r= 0.89 **B. Log-ratio Bland Altman plot comparing the CD64 expression quantification obtained from the ABxSure and the plate reader.** LoA ranged from −0.265 to +0.265 log₁₀ units. **C**. **CD64 expression quantification in the flow cytometer and ABxSure.** The solid line indicates the linear-square fit (y =218933.17x - 3.698E7), r= 0.85 **D**. **Log-ratio Bland Altman plot comparing the CD64 expression quantification obtained from the ABxSure and the flow cytometer**. LoA ranged from −0.174 to +0.174 log₁₀ units. **E**. **CD169 expression quantification in the plate reader and ABxSure.** The solid line indicates the linear-square fit (y =288.32x + 32178.43), r= 0.89 **F**. **Log-ratio Bland Altman plot comparing the CD169 expression quantification obtained from the ABxSure and the plate reader**. LoA ranged from -0.064 to +0.064 log_₁₀_ units. **G**. **CD169 expression quantification in the flow cytometer and ABxSure**. The solid line indicates the linear-square fit (y =119922.27x - 1.58E7), r= 0.89 **H**. **Log-ratio Bland Altman plot comparing the CD169 expression quantification obtained from the ABxSure and the flow cytometer**. LoA ranged from −0.255 to +0.255 log_₁₀_ units.

### 3.4 CD64-CD169 expression quantification in flow cytometer vs AB_x_Sure

Similar to the above, the blood samples were processed and evaluated using the flow cytometer to measure the CD64 and CD169 expression. The whole blood was labeled with CD64-PE and CD169-PE antibodies [In 50μL of whole blood, 3μL of CD64-PE or 1μL of CD169-PE antibody was added and incubated for 25 minutes in the dark. After incubation, 450μL of 1X lysis buffer was added and incubated for 5 minutes]. The samples were loaded in the FACS tubes in duplicates and directly used for the quantification. First, the PE calibration beads were run before each experiment and then under the same settings, samples were run where each sample was divided by the software into four distinct population according to their PE expression (Supplementary Figure S4). Granulocytes and monocytes were clearly seen with different PE expression. A total of seven tubes were prepared for each experiment-one PE calibration bead, two CD64-PE labeled blood, two CD169 labeled blood, and two unstained blood (blank).

The same sample was evaluated with the AB_x_Sure The results from both systems (flow cytometer and AB_x_Sure) were recorded and plotted in the *OriginPro* software. The x-axis represents AB_x_Sure, and the Y-axis represents the flow cytometer. The Y axis also represents the total number of PE molecules or CD64 molecules in the sample. The blank fluorescence was subtracted from the readings obtained from both the system and then plotted. The correlation between the two systems was calculated by the software and was found to be 0.85 for measuring CD64 expression and 0.89 for measuring CD169 expression in the blood samples.

Agreement between the ABxSure and flow cytometry was assessed using log–log calibration followed by log-ratio Bland–Altman analysis. Prior to calibration, large systematic differences were observed due to differences in measurement scale. After calibration, the mean log₁₀ bias was effectively reduced to zero in both datasets, indicating elimination of systematic bias. For CD64 the 95% limits of agreement ranged from −0.174 to +0.174 log₁₀ units (0.67- to 1.49-fold) (Figure 6 D, while CD169 exhibited wider limits of agreement from −0.255 to +0.255 log₁₀ units (0.56- to 1.80-fold) (Figure 6 H).

The results were also compared between plate reader and flow cytometer and plotted in the *OriginPro* software. The x-axis represents the plate reader, and the Y-axis represents the flow cytometer. The Y axis also represents the total number of PE molecules or CD64 molecules in the sample. The blank fluorescence was subtracted from the readings obtained from both systems and then plotted. The Origin software calculated the correlation between the two systems and found it to be 0.92 for measuring CD64 expression and 0.93 for measuring CD169 expression in the blood samples (Supplementary Figure S6).

## 4. Discussion

A rapid diagnostic test capable of distinguishing between bacterial and viral infections would be invaluable in healthcare settings, where timely and accurate diagnosis is critical for patient outcomes and essential for preventing AMR caused by misdiagnosis or delayed treatment. Due to overlapping symptoms of bacterial and viral infections, it is difficult to diagnose without the confirmatory microbiological culture and or PCR. This results in overprescribing of antimicrobials as a precautionary measure to mitigate the potential complications associated with severe bacterial infections. We have demonstrated a combination of biomarkers that has the potential to differentiate bacterial and viral infections by using specific immune responses. Moreover, AB_x_Sure exhibits performance comparable to the commercial plate reader and flow cytometer, a gold standard for quantifying blood cell surface proteins.

CD64 expression on the surface of neutrophils is a promising biomarker for bacterial infection and sepsis, with a sensitivity and specificity of 89.6% and 90.2%, respectively. Similarly, CD169 expression on the surface of monocytes is an efficient biomarker for detecting viral infections with sensitivity and specificity of 80% and 91%, respectively(12). CD64 and CD169 expression, along with total WBC count, will provide clinicians with a comprehensive tool for assessing a patient’s health condition.

The WBC counts from the hospital and AB_x_Sure were compared for 25 samples, yielding a correlation coefficient of 0.93. This high correlation demonstrates that AB_x_Sure is highly effective in accurately quantifying total WBCs. The Syto85 dye (λ_ex_/λ_em_ of 567/583) was used to match the excitation and emission wavelength of the phycoerythrin (λ_ex_/λ_em_ of 565/578) (fluorophore used for quantifying CD64 and CD169 expression) that is 565/578 nm. Consequently, a single optical detector employed in AB_x_Sure could be used to detect the fluorescence emission signals from the samples to quantify all three biomarkers.

Currently, the gold standard for quantification of CD64 and CD169 is flow cytometry, wherein the neutrophils and monocytes are sorted based on their size and granularity using forward and side scatter. However, the flow cytometry technique is expensive and has a bulky setup. Due to its advanced design, it usually requires trained personnel to analyze it. A study mentioned that the cost of flow cytometry-based tests can be as high as $30 per test and only 10-15 samples can be analyzed daily(17). AB_x_Sure being an automated and simpler system can overcome these challenges and provide timely results in a point-of-care setup at a much lesser cost.

CD64 and CD169 expression was quantified in three systems (plate reader, flow cytometer and AB_x_Sure) and a correlation was drawn. The correlation coefficient for total CD64 expression between the AB_x_Sure and plate reader was 0.899, while for total CD169 expression, it was 0.89. Similarly, the correlation coefficient for total CD64 expression between the AB_x_Sure and flow cytometer was 0.85, and for total CD169 expression, it was 0.89. These results indicate that AB_x_Sure is a promising device for biomarker quantification in whole blood.

Moreover, a few conclusions were drawn from the flow cytometry data analysis of the leftover blood samples used for the study. The number of CD64+ neutrophils ranged roughly between 1000 to 2000 cells, 10 times higher than CD64+ monocytes, which ranged between 100 to 200 cells. However, the number of CD64 molecules/ neutrophils was relatively low compared to CD64 molecules/ monocytes. The number of CD64 molecules/ neutrophils ranged roughly between 5000 to 8000, while CD64 molecules/ monocytes ranged roughly between 25,000 to 60,000, approximately 5 to 8 times higher (Supplementary Figure S5 A, B). This shows that the resting neutrophils, or in a no-infection state, have a slight expression of CD64 compared to monocytes, which have a constitutive expression of CD64. The CD64 expression on both neutrophils and monocytes increases in infection. The CD64 expression on neutrophils increases rapidly in cases of bacterial expression in response to interferon-γ as discussed earlier. Additionally, since the neutrophils have low expression in the no-infection state, a steep increase of >10 times is seen in cases of bacterial infections, making it a useful diagnostic marker(18). Although monocytes have high CD64 expression at all times, they show only a slight increase in the expression.

Furthermore, the neutrophil CD64 is a more specific and sensitive biomarker than monocyte CD64. When a number of neutrophils is compared with the total CD64 expression on these neutrophils, a promising linear relationship of 0.89 was observed (Supplementary Figure S5 C). As the number of neutrophils is increasing, the CD64 expression is also increasing making it a linear correlation. In contrast, the CD64 expression on monocytes is not changing much as the number increases.

Nevertheless, based on the analysis, it is clear that the total CD64 expression on WBC is also a useful infection biomarker. During infection, the number of neutrophils and their surface expression of CD64 increases significantly compared to other WBCs. A strong linear relationship was observed between the total number of CD64 molecules and the total white blood cell (WBC) count in the clinical samples (R^2^ = 0.76) (Supplementary Figure S5 E). This correlation suggests that CD64 expression scales predictably with the systemic immune response during infection. WBCs comprise ∼50 to 70% of neutrophils and ∼2 to 5% of monocytes (19). Due to this substantial difference between their numbers and the fact that monocytes have a constitutive expression of CD64, it can be proposed that total CD64 expression on WBCs can serve as a valuable biomarker for bacterial infections and the same has been demonstrated with AB_x_Sure.

It was demonstrated that AB_x_Sure is capable of quantifying cell surface biomarkers from blood, making it a promising system for infection diagnosis. Leftover blood samples were tested from patients undergoing complete blood count tests, and the system demonstrated results were comparable to those obtained with a commercial plate reader and flow cytometer. However, comparing biomarker profiles between healthy and infected patients will provide additional data for evaluating the clinical utility of AB_x_Sure.

Notwithstanding, AB_x_Sure is a compact, rapid biomarker quantification device with a total turnaround time of 20 minutes from sample addition to reading. It is an adaptable system; the users can modify sample type, cartridge reagents, label samples externally if reagent immobilization is challenging, and even customize the membrane pore size to meet specific requirements. These features make AB_x_Sure a versatile device in point-of-care diagnostics and research applications.

## Data Availability

All data produced in the present study are available upon reasonable request to the authors

## CRediT authorship contribution statement

**Neha Khaware:** Methodology, Validation, Formal analysis, Investigation, data curation, Writing-Original draft, Visualization. **Rishikant Thakur:** Mechanical design, Writing-Review and Editing and fabrication. **Kartik Kachhawa**: Fabrication and Software. **Prabhu Balasubramanian:** Fabrication, Software, Data curation. **Vivekanandan Perumal:** Writing-Review and Editing, Supervision, Funding acquisition. **Ravikrishnan Elangovan:** Conceptualization, Methodology, Resources, Writing-Review and Editing, Supervision, Project administration, Funding acquisition.

## Acknowledgments

This study is completed as part of the DOSA Project (Diagnostics for One Health and User Driven Solutions for AMR, www.dosa-diagnostics.org) funded by the Government of India’s Department of Biotechnology (grant number: BT/IN/Indo-UK/AMR/03/). The Ministry of Education (MoE), India under the Grand Challenge Project scheme (MI1798; received by RE and VP) of the Industrial Research & Development Unit, Indian Institute of Technology Delhi.

## Appendix A. Supplementary data

