## Supplemental data for "AB_x_Sure for differentiating bacterial from viral Infection"

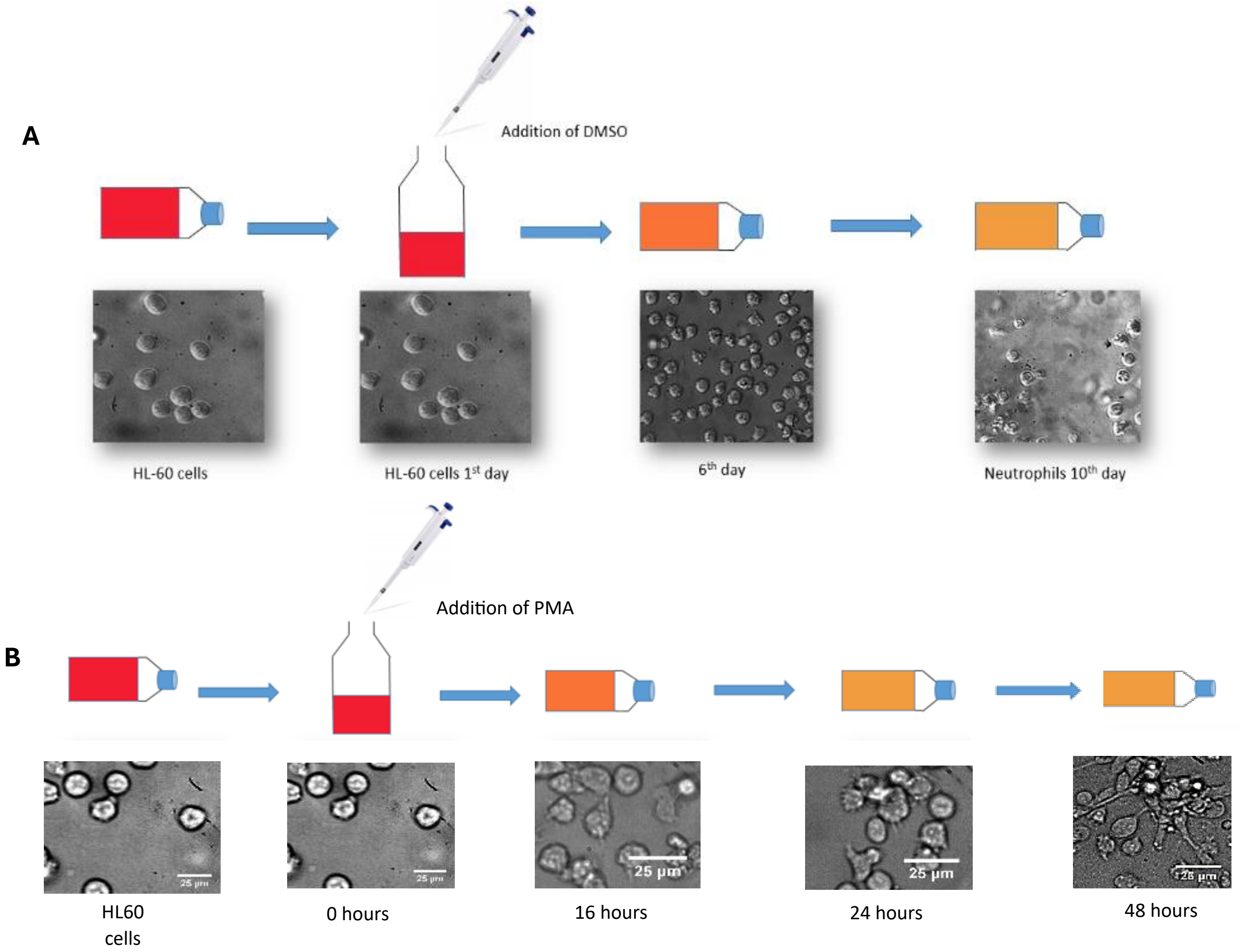


Supplementary Figure S1: **A. Differentiation of HL60 cells into Neutrophils using DMSO.** The HL60 cells were incubated with 1.3% DMSO for 8 days; the media was changed every three days. **B. Differentiation of HL60 cells into Monocyte using PMA.** The HL60 cells were incubated with 2ng/mL of phorbol myristate acetate (PMA) for 2 days. All incubations were performed in the CO2 incubator at 37 °C and 5% $CO_2$.

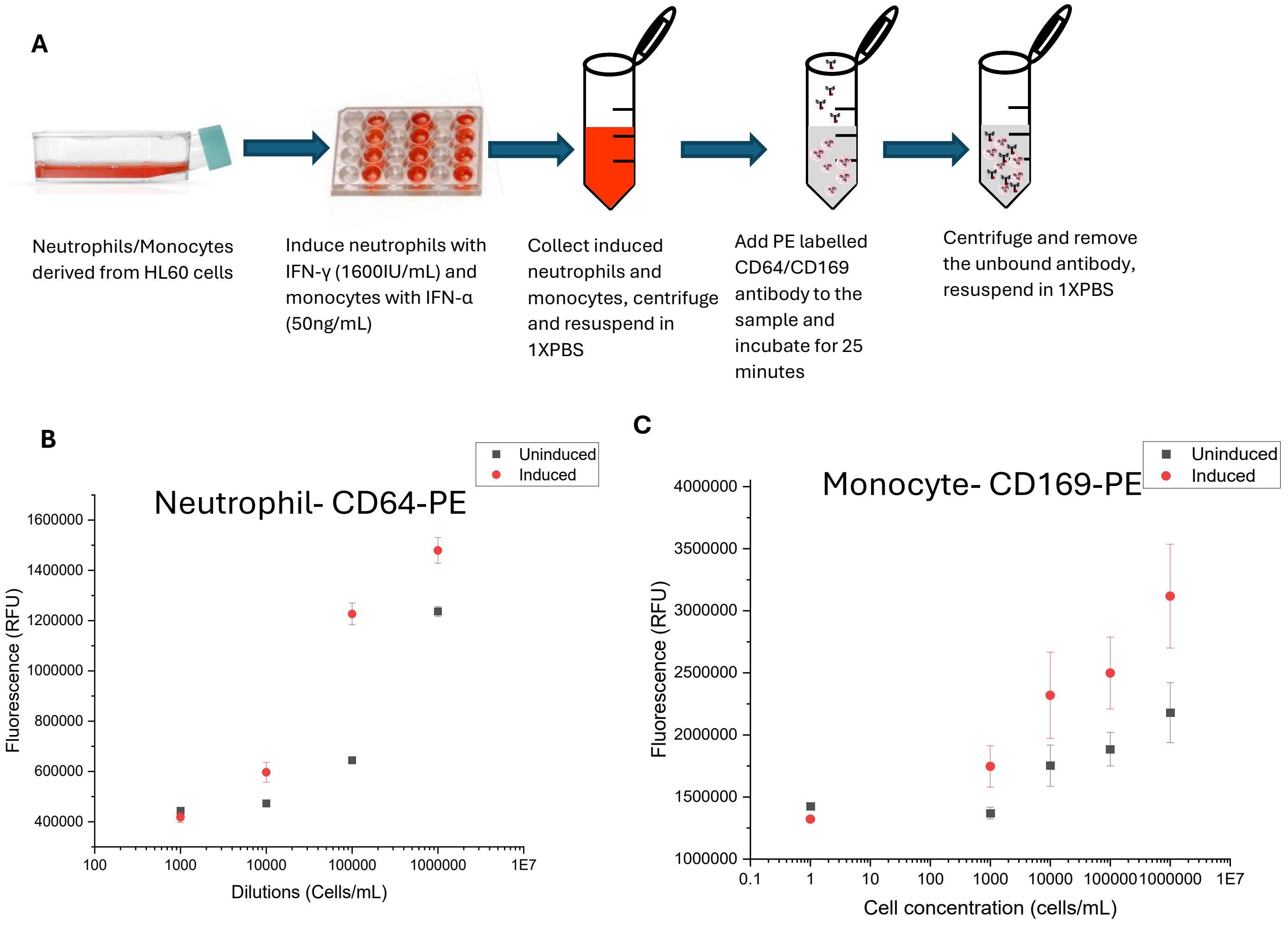


Supplementary Figure S2: **A. Protocol for neutrophil induction by IFN-γ/ IFN-α and CD64/ CD169 expression quantification in plate reader. B. CD64-PE and quantification in Neutrophils derived from HL60 cells (n=3).** *Error bars represent standard deviation*. **C. CD169-PE quantification in monocytes derived from HL60 cells (n=3).** *Error bars represent standard deviation.*

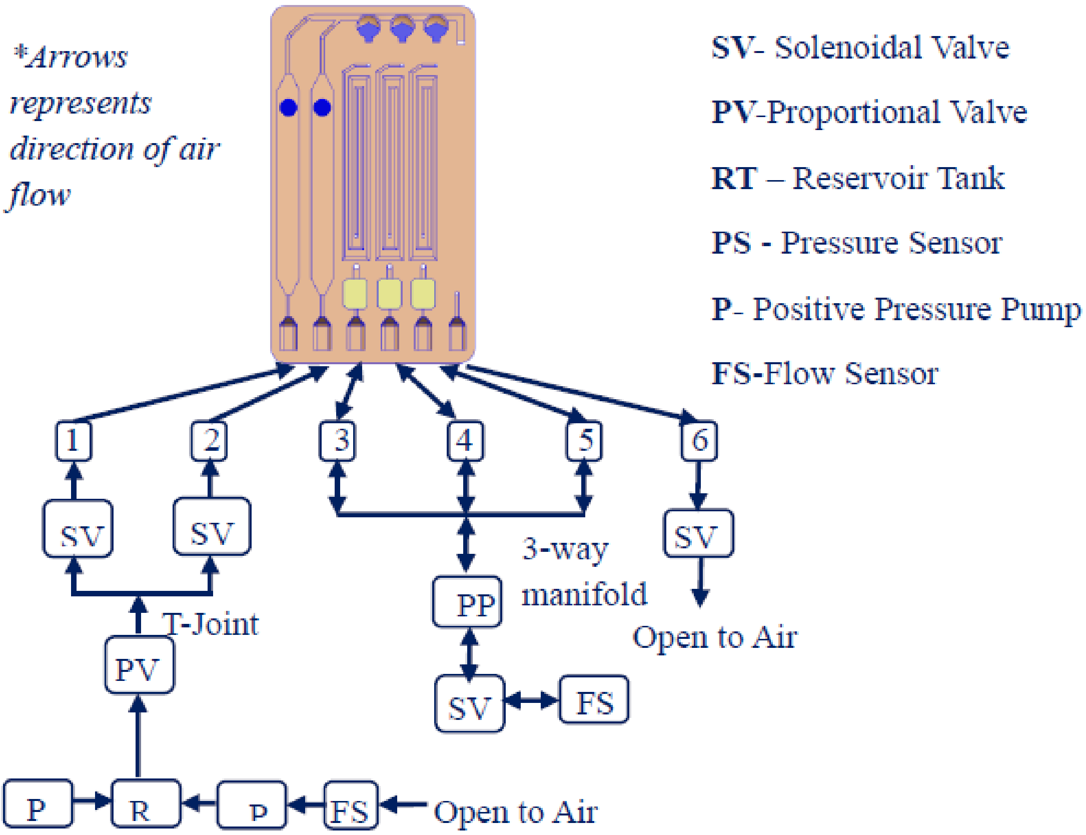


Supplementary Figure S3: **Schematic representation of the pneumatic control in the AB$_x$Sure**

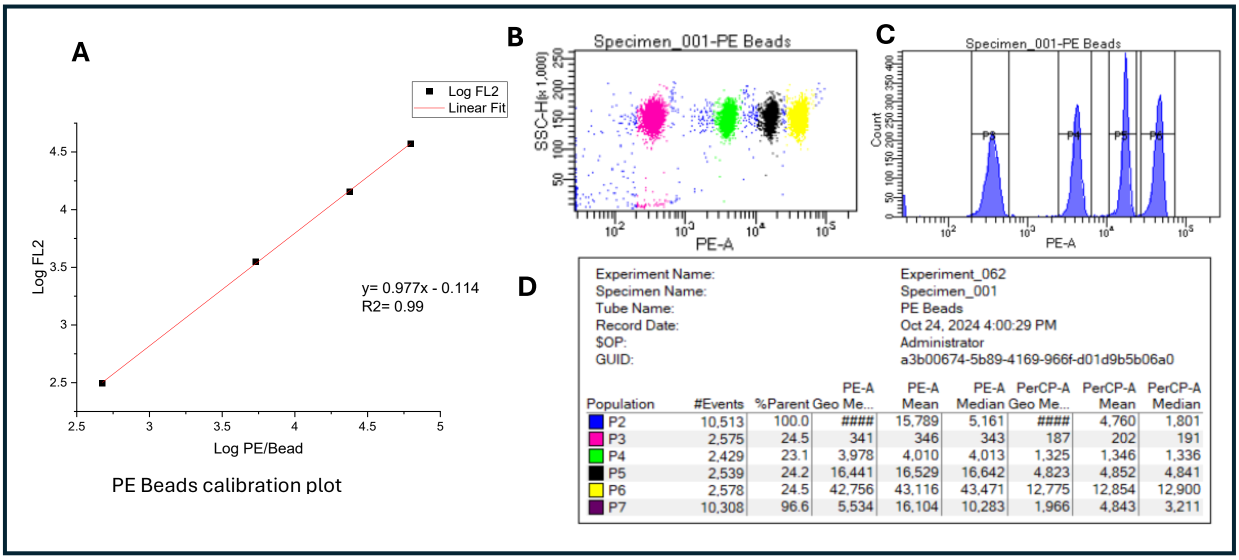


| | |
|---|---|
| Experiment Name: | Experiment_062 |
| Specimen Name: | Specimen_001 |
| Tube Name: | PE Beads |
| Record Date: | Oct 24, 2024 4:00:29 PM |
| $OP: | Administrator |
| GUID: | a3b00674-5b89-4169-966f-d01d9b5b06a0 |

| Population | #Events | %Parent | PE-A Geo Me... | PE-A Mean | PE-A Median | PerCP-A Geo Me... | PerCP-A Mean | PerCP-A Median |
|---|---|---|---|---|---|---|---|---|
| P2 | 10,513 | 100.0 | #### | 15,789 | 5,161 | #### | 4,760 | 1,801 |
| P3 | 2,575 | 24.5 | 341 | 346 | 343 | 187 | 202 | 191 |
| P4 | 2,429 | 23.1 | 3,978 | 4,010 | 4,013 | 1,325 | 1,346 | 1,336 |
| P5 | 2,539 | 24.2 | 16,441 | 16,529 | 16,642 | 4,823 | 4,852 | 4,841 |
| P6 | 2,578 | 24.5 | 42,756 | 43,116 | 43,471 | 12,775 | 12,854 | 12,900 |
| P7 | 10,308 | 96.6 | 5,534 | 16,104 | 10,283 | 1,966 | 4,843 | 3,211 |

Supplementary Figure S4: **Flow cytometry protocol with BD Quantibrite calibration kit**. **A**. PE Beads calibration plot, **B**. Separation of four distinct population (with different number of bound PE molecules) in the Flow software(Pink=nearly no expression, Green= low expression, Black= medium expression and yellow= high expression), **C**. P3 to P6 are four populations. The count indicates the number of beads with increasing number of PE molecules. **D**. Events indicate how many times a particular bead population was detected by the software, geometric mean in 3$^{rd}$ column indicates number of PE molecules per bead

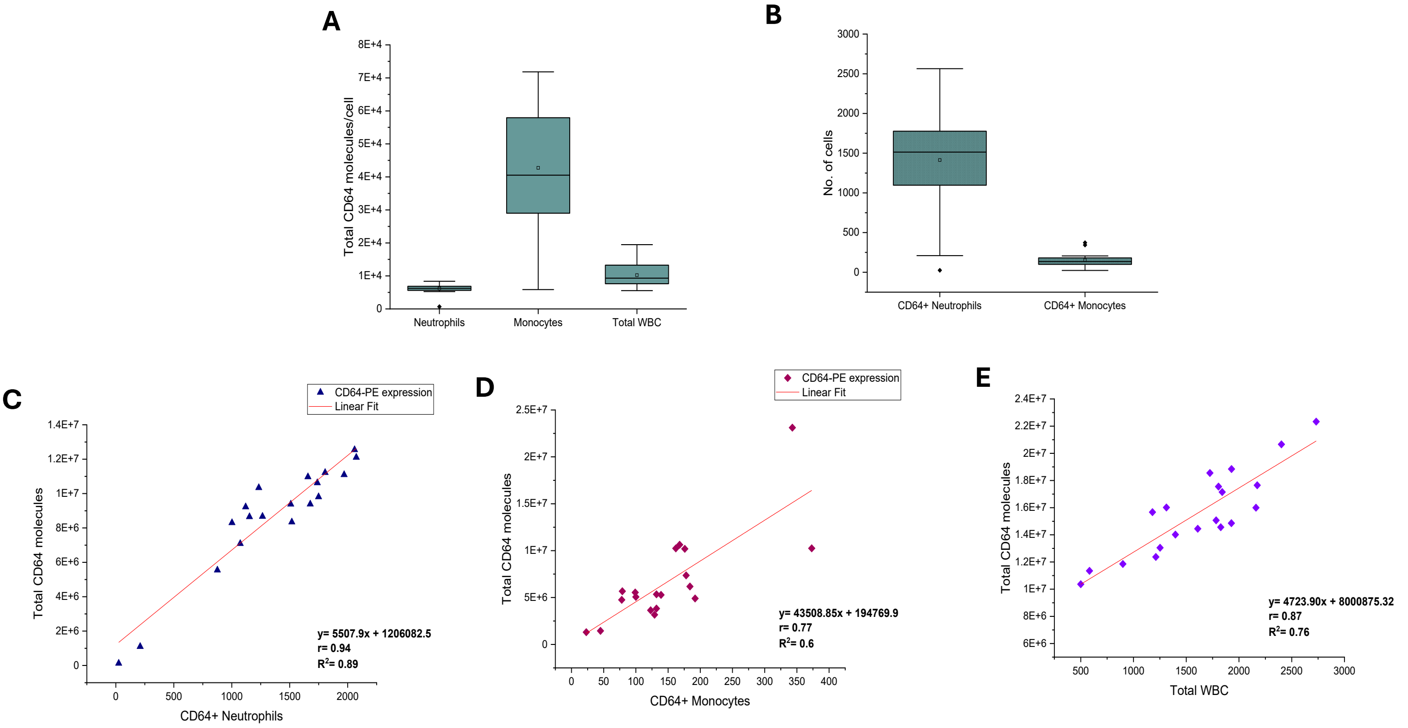


Supplementary Figure S5: **Flow cytometry data analysis**. **A. Total CD64 molecules/cells in neutrophils, monocytes and total WBC**, **B. CD64+ neutrophils vs CD64+ monocytes**, **C. Correlation between no. of neutrophils and total CD64 expression**. The solid line indicates the linear-square fit (y = 5507.9x + 1206082.5) $R^2$= 0.89, r= 0.84 **D. Correlation between no. of monocytes and total CD64 expression**. The solid line indicates the linear-square fit (y = 43508.85x + 194769.9) $R^2$= 0.6, r= 0.77 **E. Correlation between total WBC vs total CD64 molecules expressed**. The solid line indicates the linear-square fit (y = 4723.90x + 8000875.32) $R^2$= 0.76, r= 0.87

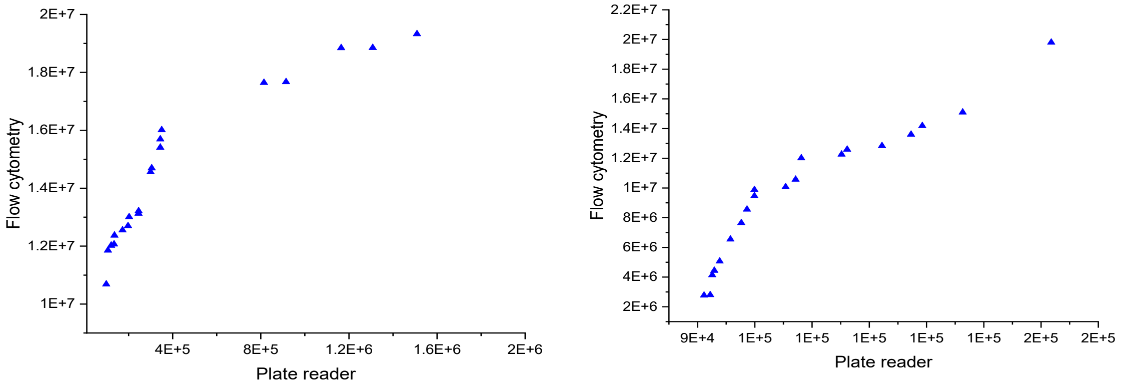


Supplementary Figure S6: **CD64 (A) and CD169 (B) expression quantification in the plate reader (VictorNivo, Revvity) vs flow cytometer (BD Fortessa X20)**
